# US Public Opinion as to Whether “The Pandemic is Over,” September to October 2022

**DOI:** 10.1101/2022.12.23.22283899

**Authors:** Mark É Czeisler, Matthew D Weaver, Rashon I Lane, Shantha MW Rajaratnam, Mark E Howard, Charles A Czeisler

## Abstract

**Importance:** US public health guidance has increasingly shifted responsibility for actions to minimize ongoing impacts of COVID-19 onto individuals. During September to October 2022, the World Health Organization continued to characterize COVID-19 as a pandemic. Yet, public perceptions of the pandemic status of COVID-19 and its associations with COVID-19-related behaviors were unknown.

**Objective:** To assess US public opinion on the characterization of COVID-19 as a pandemic.

**Design, Setting, and Participants:** The COVID-19 Outbreak Public Evaluation (COPE) Initiative internet-based surveys, administered to 4985 US adults during September to October 2022. Demographic quota sampling and survey weighting were employed to improve sample representativeness of the US population by age, sex, and combined race and ethnicity.

**Exposures:** The COVID-19 pandemic.

**Main Outcomes and Measures:** Response to the statement, “The pandemic is over.”‘ Response options included Strongly agree, Somewhat agree, Neutral, Somewhat disagree, and Strongly disagree.

**Results:** Overall, 5015 US adults completed The COPE Initiative surveys (response rate, 56.2%), and 4985 (99.4%) provided complete information for all analyzed variables and were included in this analysis. Only 1657 (33.2%) respondents agreed with the statement “the pandemic is over,” while 2141 (43.0%) disagreed and the remaining 1187 (23.8%) were neutral about the statement. Agreement that the pandemic was over was most strongly associated with having received fewer COVID-19 vaccines, lesser concern about SARS-CoV-2 variant viruses, and less frequent engagement in COVID- 19 preventive behaviors, such as mask usage in public spaces, as well as increasingly conservative political ideology, roles as unpaid caregivers of both children and adults, younger age, male sex, and significant disabilities.

**Conclusions and Relevance:** As of September to October 2022, US public opinion was mixed on the characterization of COVID-19 as a pandemic. Belief the pandemic was over was associated with less frequent engagement in COVID-19 preventive\behaviors, highlighting the important role of public health communication. Demographic groups to prioritize tailored public health messaging about the pandemic status were identified. Continued assessment of public perceptions about the state of the pandemic is warranted entering Year 4 of the COVID-19 pandemic.

**Key Points:** 

**Question:** As of September to October 2022, what was US public opinion as to whether COVID-19 remained a pandemic?

**Findings:** In this demographically representative survey study of 4985 US adults, only 1 in 3 respondents agreed with the statement “the pandemic is over;” 43% of adults disagreed. Agreement that the pandemic was over was associated with less engagement in COVID-19 preventive behaviors and more political conservatism.

**Meaning:** As of September to October 2022, US public opinion was divided regarding the status of COVID-19 as a pandemic and is associated with COVID-19-related behaviors, underscoring important public health and policy implications of this designation.

## Introduction

On March 11, 2020, the Director-General of the World Health Organization (WHO) officially characterized the coronavirus disease 2019 (COVID-19) outbreak as a global pandemic.^**1**^ Two days later, the President of the United States proclaimed that COVID-19 constituted a national emergency.^**2**^ In the initial pandemic months, absent vaccines to prevent SARS-CoV-2 transmission or COVID-19 medical treatments, community COVID-19 prevention measures were implemented. Stay-at-home orders and nonessential business closures were executed worldwide for the first time in a century. Early support for and adherence with these nonpharmaceutical interventions^**3–5**^ waned with pressures to reopen economies and resume in-person activities.^**6–8**^

Despite a better understanding of SARS-CoV-2 transmission dynamics, the rapid development of COVID-19 vaccines, and considerable advances in COVID-19 medical treatments, COVID-19 has remained a major cause of global morbidity and mortality in many areas worldwide, with the US among the most adversely impacted nations. Indeed, COVID-19-involved mortality and excess all-cause mortality were high in the US relative to comparator countries during the initial pandemic wave through September 2020,^**9**^ and throughout the Delta and winter Omicron waves during June 2021 through March 2022.^**10**^ The prolonged severity of the pandemic is due in part to the emergence of immune-evasive coronavirus variant viruses and incomplete vaccine uptake, which have thwarted achievement of sufficient population-level immunity to reduce sustained transmission. The prolonged waves of coronavirus infections and associated public health containment measures have induced pandemic fatigue and pandemic-related political polarization, lowering adherence with nonpharmaceutical interventions such as masking. Throughout the pandemic, longstanding and worsening social inequities have predisposed historically marginalized and oppressed groups to both COVID-19-involved and non-COVID-19-involved adverse health outcomes^**11,12**^ partly due to a higher prevalence of baseline chronic medical conditions,^**13**^ more frequent exposure to infections, and less access to healthcare prevention and treatment services.^**14**^

As of mid-September 2022, 18 months after the declaration of the pandemic by the WHO, there had been approximately 6.5 million COVID-19-involved deaths and 600 million SARS-CoV-2 infections documented globally, with over 1 million deaths and nearing 100 million infections occurring in the United States.^**15**^ The approximately 400,000-500,000 weekly confirmed cases documented in mid-September 2022 were higher than the 200,000-250,000 weekly confirmed cases documented in mid-September 2020 and lower than the 1 million weekly confirmed cases documented in mid-September 2021, during the wave of the highly transmissible Delta (B.1.617.2) variant.^**15**^ While comparisons are limited by the variable number and types of tests performed at different stages of the pandemic (eg, approximate weekly COVID-19 Nucleic Acid Amplification Tests reported to the CDC in mid-September 2020 [6.5 to 7 million]; 2021 [11.5-12 million]; and 2022 [3.3-3.5 million]),^**16**^ the persistence of hundreds of thousands of detected infections despite reduced reported testing serves as evidence of sustained SARS-CoV-2 transmission in the US.

Additionally, though prevalence estimates vary widely due to differing definitions of post-acute sequelae of SARS-CoV-2 infection—also known as Long COVID and Post-COVID Conditions—abundant emerging evidence points to long-term sequelae for a substantial percentage of people following the acute phase of infection, including pulmonary, cardiovascular, renal, hepatic, neurologic, and mental health sequelae, as well as functional mobility impairments and general and constitutional symptoms.^**17–19**^

Without an official announcement from the World Health Organization or US Centers for Disease Control and Prevention (CDC), on September 18, 2022, the President of the United States twice declared “the pandemic is over” on an American television news magazine broadcast. On September 20, 2022, the President addressed the controversial comments at the United Nations General Assembly, clarifying he had intended to convey that the US was in a different place due to vaccines and post-infectious immunity. At that same Assembly, the WHO Director-General stated that the pandemic was not over, and on September 22, 2022, the CDC Director neither directly agreed nor disagreed with the president’s assertion.

Public perceptions about the pandemic are consequential. A recent study found that only 15% of surveyed adults living in areas with high COVID-19 transmission during May through July 2022 realized that COVID-19 transmission in their local area was high at that time.^**20**^ Lower perceived local community transmission levels were associated with having received fewer COVID-19 vaccines, lower concern about SARS-CoV-2 variant viruses, and less frequent engagement in COVID-19 preventive behaviors. Moreover, respondents indicated that they would change their behaviors in response to levels of local COVID-19 transmission, highlighting that keeping the public fully informed about the state of the pandemic, which is an essential tenant of public health,^**21**^ could influence engagement in behaviors to reduce transmission of SARS-CoV-2.

We therefore sought to estimate the prevalence of US adults who agreed with, felt neutral about, and disagreed with the statement, “the pandemic is over,” and to characterize the association between agreement with the pandemic-over statement and engagement in behaviors to reduce SARS-CoV-2 infections and prevent severe COVID-19 illness. Additionally, we endeavored to characterize demographic associations with agreement with the pandemic-over statement to identify groups to prioritize tailored public health communication strategies.

## Methods

### Setting and Participants

During September 26 to October 15, 2022 (ie, approximately 1 to 3 weeks after the President’s “the pandemic is over” statement), US adults aged ≥ 18 years completed internet-based surveys through Qualtrics for The COVID-19 Outbreak Public Evaluation (COPE) Initiative (www.thecopeinitiative.org). Demographic quota sampling and survey weighting were employed to improve sample representativeness of the US population by age, sex, and combined race and ethnicity. Respondents were not informed of survey topics prior to commencement.

### Key Definitions

#### Main Outcome

Respondents were asked, ‘Select your response to the statement, “The pandemic is over.”‘ Response options included Strongly agree, Somewhat agree, Neutral, Somewhat disagree, and Strongly disagree.

#### Demographic Characteristics

Demographic variables included in this analysis were sex at birth, age group in years, race, ethnicity, highest education attainment, experience with disability, self-rated health, roles as unpaid caregivers, state of residence (categorized as 4 US Census Bureau-designated regions), and political ideology. Response options and analytic categorizations are detailed in the Supplement.

#### Pandemic-related Characteristics

Pandemic-related variables included in this analysis were frequency of mask usage in public and avoidance of large gatherings, COVID-19 vaccine doses received, and concern about the health effects of coronavirus variants circulating at the time respondents were surveyed. Response options and analytic categorizations are detailed in the Supplement.

#### Statistical Analyses

Rounded, weighted counts and percentages were used to describe the sample demographics and outcomes. Multivariable Poisson regressions with robust standard errors were used to estimate adjusted prevalence ratios (aPRs) and 95% CIs for agreement (somewhat or strongly) that the pandemic was over when surveyed, adjusted for the following: sex, age, combined race and ethnicity, education attainment, and unpaid caregiver roles. Separate models were run for collinear variables: disability and self-rated health, Census region and political ideology. Calculations were performed in Python (V3.7.8) and R (V4.2.0). Statistical significance was defined as 95% CIs excluding the null value.

#### Study Approval and Consent

The Monash University Human Research Ethics Committee approved the study protocol; participants provided informed consent electronically. Detailed Methods are in the Supplement.

## Results

Overall, 5015 of 8930 invited eligible participants (56.2%) completed The COPE Initiative surveys; 4985 of these respondents (99.4%) met secondary screening criteria and were analyzed (Figure 1). Overall, 1657 (33.2%) respondents indicated that they agreed with the statement “The pandemic is over” (661 [13.3%] strongly agreed; 996 [20.0%] somewhat agreed); 2141 (43.0%) indicated that they disagreed (954 [19.1%] strongly disagreed; 1188 [23.8%] somewhat disagreed); and 1187 (23.8%) provided a neutral response (Table 1).

**Table 1.** Response to the Statement “The Pandemic is Over” Among US Adults, Overall and by Demographic Characteristics, September to October 2022.

|  | All respondents | Strongly disagree | Somewhat disagree | Neutral | Somewhat agree | Strongly agree | P |
| --- | --- | --- | --- | --- | --- | --- | --- |
|  | No. (%) | No. (%) | No. (%) | No. (%) | No. (%) | No. (%) |  |
| **All respondents** | **4985 (100)** | **954 (19.1)** | **1188 (23.8)** | **1187 (23.8)** | **996 (20.0)** | **661 (13.3)** |  |
| **Sex** |  |  |  |  |  |  |  |
| Female | 2539 (50.9) | 577 (22.7) | 639 (25.2) | 639 (25.2) | 450 (17.7) | 234 (9.2) | <.001 |
| Male | 2446 (49.1) | 376 (15.4) | 549 (22.4) | 548 (22.4) | 546 (22.3) | 427 (17.5) |  |
| **Age group, years** |  |  |  |  |  |  |  |
| 18 to 24 | 581 (11.6) | 85 (14.6) | 118 (20.3) | 176 (30.2) | 132 (22.8) | 70 (12.1) | <.001 |
| 25 to 44 | 1716 (34.4) | 267 (15.6) | 305 (17.8) | 464 (27.1) | 380 (22.2) | 299 (17.4) |  |
| 45 to 64 | 1611 (32.3) | 345 (21.4) | 401 (24.9) | 349 (21.7) | 313 (19.4) | 203 (12.6) |  |
| 65 and over | 1078 (21.6) | 257 (23.8) | 363 (33.7) | 198 (18.3) | 171 (15.9) | 89 (8.3) |  |
| **Combined race and ethnicity<sup>a</sup>** |  |  |  |  |  |  |  |
| Asian, non-Hispanic | 303 (6.1) | 61 (20.1) | 86 (28.2) | 69 (22.8) | 61 (20.1) | 26 (8.7) | <.001 |
| Black, non-Hispanic | 612 (12.3) | 160 (26.1) | 148 (24.2) | 158 (25.8) | 99 (16.2) | 48 (7.8) |  |
| Hispanic or Latino, any race or races | 844 (16.9) | 170 (20.2) | 190 (22.5) | 207 (24.5) | 161 (19.1) | 116 (13.7) |  |
| Other race or races, non-Hispanic | 129 (2.6) | 26 (19.8) | 20 (15.7) | 52 (40.0) | 15 (12.0) | 16 (12.4) |  |
| White, non-Hispanic | 3097 (62.1) | 537 (17.3) | 744 (24.0) | 702 (22.6) | 659 (21.3) | 455 (14.7) |  |
| **Highest education attainment** |  |  |  |  |  |  |  |
| High school degree or less | 1224 (24.5) | 272 (22.2) | 226 (18.5) | 403 (32.9) | 206 (16.9) | 117 (9.5) | <.001 |
| College or some college | 2882 (57.8) | 540 (18.7) | 772 (26.8) | 635 (22.0) | 580 (20.1) | 356 (12.3) |  |
| More than bachelor's degree | 879 (17.6) | 142 (16.1) | 190 (21.6) | 149 (17.0) | 210 (23.9) | 189 (21.5) |  |
| **Experience with disability<sup>b</sup>** |  |  |  |  |  |  |  |
| No difficulty with tasks | 1954 (39.2) | 364 (18.6) | 471 (24.1) | 491 (25.1) | 377 (19.3) | 252 (12.9) | <.001 |
| Some difficulty with tasks | 1678 (33.7) | 366 (21.8) | 484 (28.8) | 338 (20.2) | 316 (18.8) | 174 (10.4) |  |
| A lot of difficulty with tasks | 974 (19.5) | 178 (18.3) | 190 (19.5) | 254 (26.1) | 198 (20.3) | 154 (15.8) |  |
| Cannot do some tasks at all | 379 (7.6) | 46 (12.0) | 43 (11.3) | 104 (27.3) | 105 (27.7) | 82 (21.6) |  |
| **Self-rated health** |  |  |  |  |  |  |  |
| Excellent | 846 (17.0) | 106 (12.6) | 110 (13.0) | 185 (21.9) | 169 (20.0) | 276 (32.6) | <.001 |
| Very good | 1561 (31.3) | 262 (16.8) | 360 (23.1) | 355 (22.7) | 403 (25.8) | 181 (11.6) |  |
| Good | 1662 (33.3) | 353 (21.2) | 433 (26.1) | 479 (28.8) | 272 (16.4) | 124 (7.5) |  |
| Fair | 761 (15.3) | 184 (24.2) | 242 (31.8) | 143 (18.8) | 124 (16.2) | 68 (9.0) |  |
| Poor | 154 (3.1) | 48 (31.3) | 43 (27.6) | 25 (16.1) | 27 (17.7) | 11 (7.3) |  |
| **Unpaid caregiver roles** |  |  |  |  |  |  |  |
| None | 3412 (68.4) | 686 (20.1) | 873 (25.6) | 839 (24.6) | 628 (18.4) | 387 (11.3) | <.001 |
| Unpaid caregiver of adults only | 540 (10.8) | 140 (25.9) | 119 (22.0) | 115 (21.3) | 102 (18.9) | 64 (11.8) |  |
| Unpaid caregiver of children only | 424 (8.5) | 58 (13.7) | 121 (28.6) | 95 (22.4) | 109 (25.6) | 41 (9.7) |  |
| Unpaid caregiver of both age groups | 609 (12.2) | 70 (11.5) | 74 (12.2) | 138 (22.6) | 157 (25.8) | 170 (27.8) |  |
| **US Census region<sup>c</sup>** |  |  |  |  |  |  |  |
| Northeast | 935 (18.8) | 175 (18.7) | 207 (22.1) | 237 (25.3) | 182 (19.5) | 134 (14.3) | >.99 |
| Midwest | 980 (19.7) | 166 (16.9) | 238 (24.3) | 221 (22.5) | 217 (22.1) | 139 (14.2) |  |
| South | 1863 (37.4) | 365 (19.6) | 434 (23.3) | 460 (24.7) | 356 (19.1) | 248 (13.3) |  |
| West | 1206 (24.2) | 248 (20.6) | 307 (25.5) | 269 (22.3) | 241 (20.0) | 140 (11.6) |  |
| **Political ideology<sup>d</sup>** |  |  |  |  |  |  |  |
| Very liberal | 797 (16.0) | 190 (23.9) | 190 (23.8) | 144 (18.0) | 158 (19.8) | 115 (14.4) | <.001 |
| Slightly liberal | 869 (17.4) | 214 (24.7) | 268 (30.8) | 150 (17.3) | 173 (19.9) | 63 (7.3) |  |
| Neither liberal nor conservative | 1404 (28.2) | 264 (18.8) | 361 (25.7) | 453 (32.2) | 231 (16.4) | 96 (6.8) |  |
| Slightly conservative | 848 (17.0) | 117 (13.8) | 210 (24.7) | 173 (20.3) | 226 (26.6) | 123 (14.5) |  |
| Very conservative | 824 (16.5) | 113 (13.7) | 117 (14.2) | 169 (20.5) | 182 (22.0) | 244 (29.6) |  |

Abbreviations: COVID-19, coronavirus disease 2019
<sup>a</sup> Includes “American Indian or Alaskan Native,” “Native Hawaiian or other Pacific Islander,” and “Other.”
<sup>b</sup> Based on the Washington Group - Short Set of Questions on Disability. Includes difficulty seeing, hearing, walking or climbing steps, remembering or concentrating, with self-care, and communicating.
<sup>c</sup> A single respondent living in Puerto Rico is not included in the US Census Region rows.
<sup>d</sup> Respondents who preferred not to answer with their political ideology (n = 243) are not included in the Political ideology tables.

**Figure 1.**
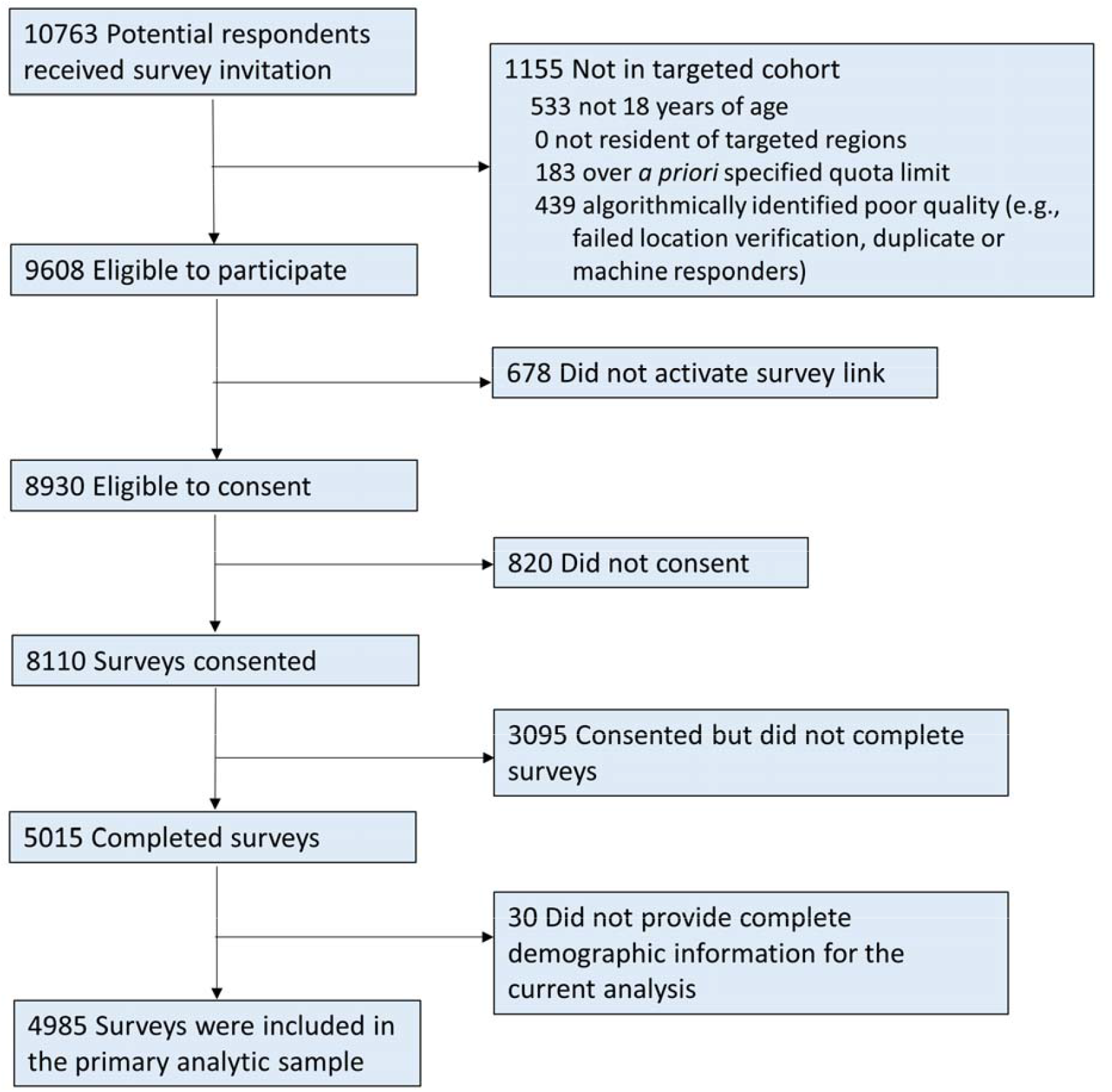
Flow of The COPE Initiative Survey Respondents, September to October 2022.

Responses to the pandemic-over statement also varied by demographic characteristics (Table 1). Groups in which more than 40% of respondents agreed the pandemic was over included respondents who were unpaid caregivers of both children and adults (376 of 609 respondents [53.6%]); those who reported their self-rated health as excellent (445 of 846 respondents [52.6%]); those who identified their political ideology as very conservative (425 of 824 respondents [51.6%]) or slightly conservative (349 of 848 respondents [41.2%]); those who identified as having disabilities such that they could not do some tasks at all (187 of 379 respondents [49.3%]); and those who had education attainment of more than a bachelor’s degree (399 of 879 respondents [45.3%]). Conversely, groups in which less than 25% of respondents agreed the pandemic was over included respondents who identified their political ideology as neither liberal nor conservative (327 of 1404 respondents [23.3%]); those who identified their combined race and ethnicity of Black, non-Hispanic (147 of 612 respondents [24.0%]) and other race or multiple races, non-Hispanic (31 of 129 respondents [24.4%]); those who were of age 65 years and over (260 of 1078 respondents [24.1%]); and those who self-rated their health as poor (39 of 154 respondents [25.0%]).

Responses to the pandemic-over statement also varied by COVID-19-related attitudes and behaviors (Table 2). Groups in which more than 40% of respondents agreed the pandemic was over included respondents who in the 2 weeks preceding the survey never avoided large gatherings (686 of 1707 respondents [40.2%]) or never wore a mask in public spaces (702 of 1537 respondents [45.6%]); those who had received only one COVID-19 vaccine dose (163 of 385 respondents [42.5%]); and those who were not at all concerned (487 of 934 respondents [52.2%]) or somewhat unconcerned (218 of 495 respondents [44.0%]) about current SARS-CoV-2 variant viruses. Conversely, groups in which less than 25% of respondents agreed the pandemic was over included respondents who had received four or more COVID-19 vaccine doses (155 of 852 respondents [18.2%]); those who in the 2 weeks preceding the survey had sometimes worn a face mask in public (208 of 859 respondents [24.2%]) or not gone to public places (33 of 150 respondents [21.8%]); and those who were somewhat concerned (364 of 1484 respondents [24.5%]) or neutral (293 of 1209 respondents [24.2%]) about current SARS-CoV-2 variant viruses.

**Table 2.**
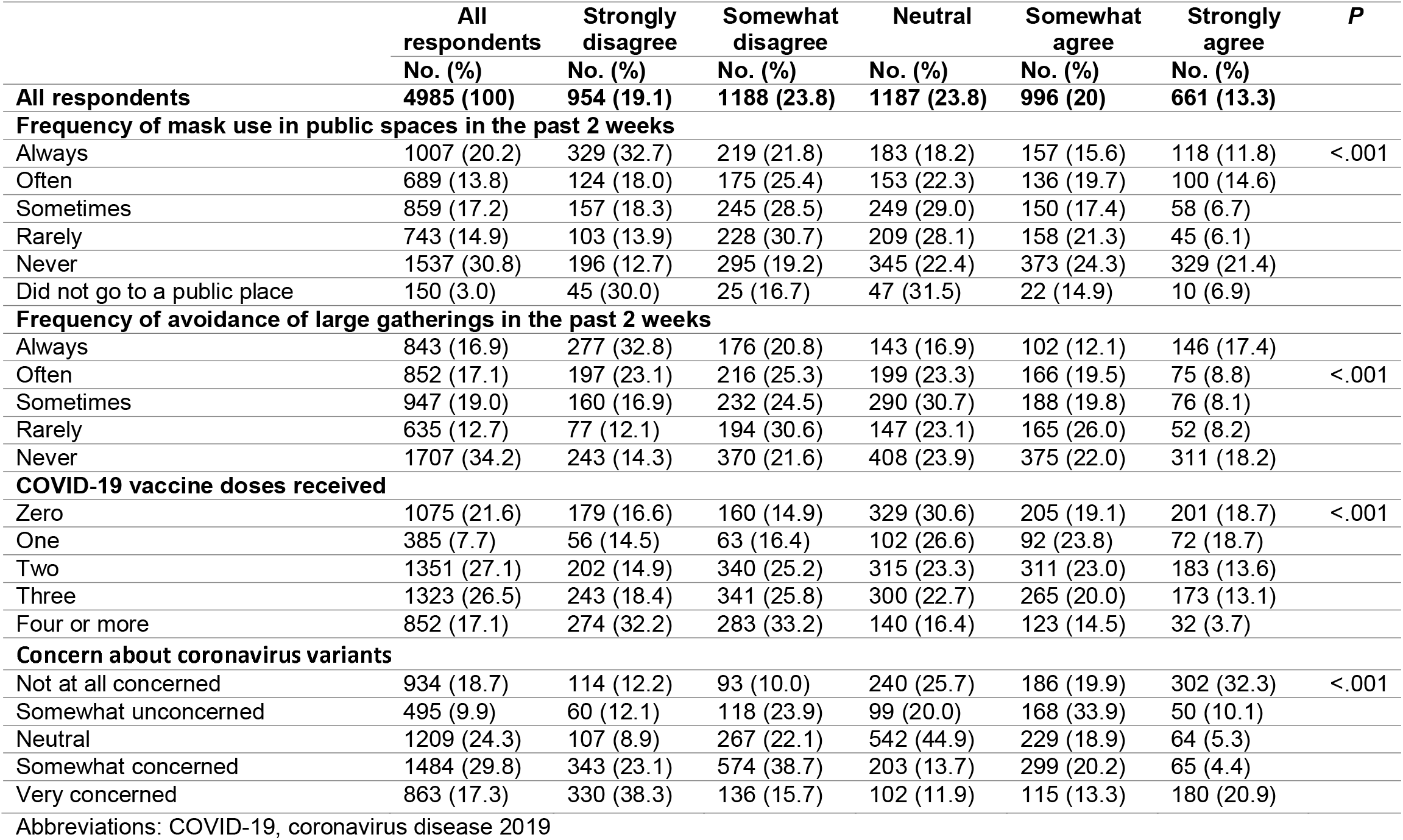
Response to the Statement “The Pandemic is Over” Among US Adults, Overall and by COVID-19-Related Characteristics, September to October 2022.

The multivariable model adjusting for demographic variables only revealed higher odds of agreement with the pandemic-over statement among respondents with conservative vs liberal political ideology (eg, very conservative vs very liberal, aOR, 2.6 [95% CI, 2.1 to 3.3]); those with roles as unpaid caregivers of both children and adults vs those without caregiver roles (aOR, 2.1 [95% CI, 1.7 to 2.5]); those of male vs female sex (aOR, 1.6 [95% CI, 1.4 to 1.9]); those of younger vs older age (eg, 18 to 24 vs 65 or more years, aOR, 1.8 [95% CI, 1.3 to 2.5]); and those with disabilities such that they could not do some tasks at all vs without disabilities (aOR, 1.5 [95% CI, 1.1 to 1.9]) (Figure 2). Lower odds of agreement were found for respondents with worse self-rated health (eg, poor vs excellent, aOR, 0.4 [95% CI, 0.3 to 0.7]); those with lower education attainment (eg, high school degree or less vs more than a bachelor’s degree, aOR, 0.5 [95% CI, 0.4 to 0.7]); and those who identified their combined race and ethnicity as something other than non-Hispanic White (eg, non-Hispanic Black, aOR, 0.6 [0.5 to 0.7]).

**Figure 2.**
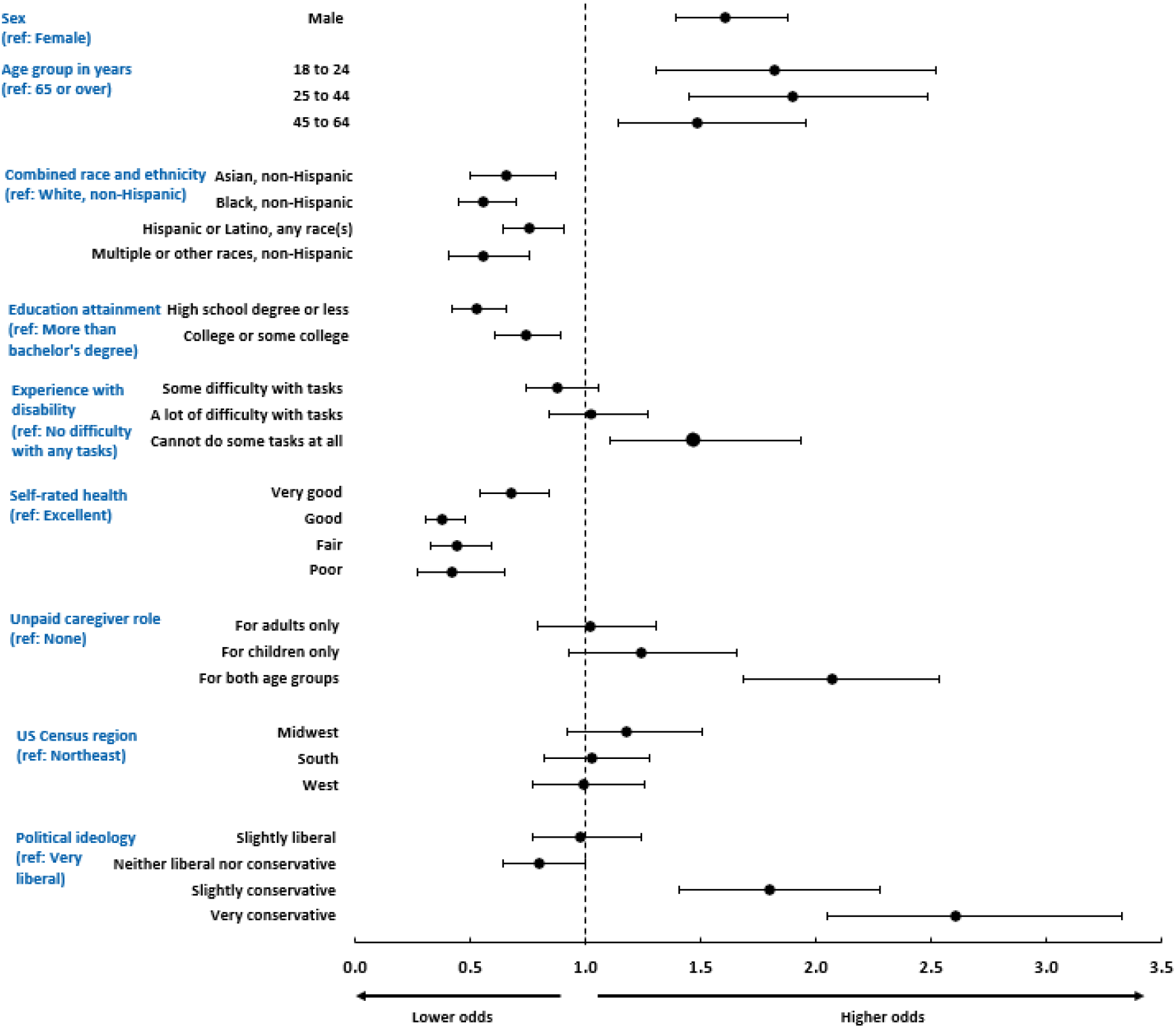
Adjusted Odds Ratios of Having Agreed that the Pandemic was Over by Demographic Characteristics, September to October 2022.

Multivariable models adjusting for demographic variables plus individual COVID-19-related variables revealed higher odds of agreement with the pandemic-over statement among respondents who in the 2 weeks preceding the survey never vs always wore masks in public spaces (aOR, 3.0 [95% CI, 2.4 to 3.7]); those who had rarely or never vs always avoided large gatherings (eg, never, aOR, 2.2 [95% CI, 1.7 to 2.7]) or had received fewer vs more COVID-19 vaccine doses (e.g., zero vs four or more doses, aOR, 3.4 [95% CI, 2.5 to 4.6]); and who were not at all concerned or somewhat unconcerned vs very concerned about SARS-CoV-2 variant viruses (eg, not at all concerned, aOR, 3.2 [95% CI, 2.5 to 4.0]) (Figure 3).

**Figure 3.**
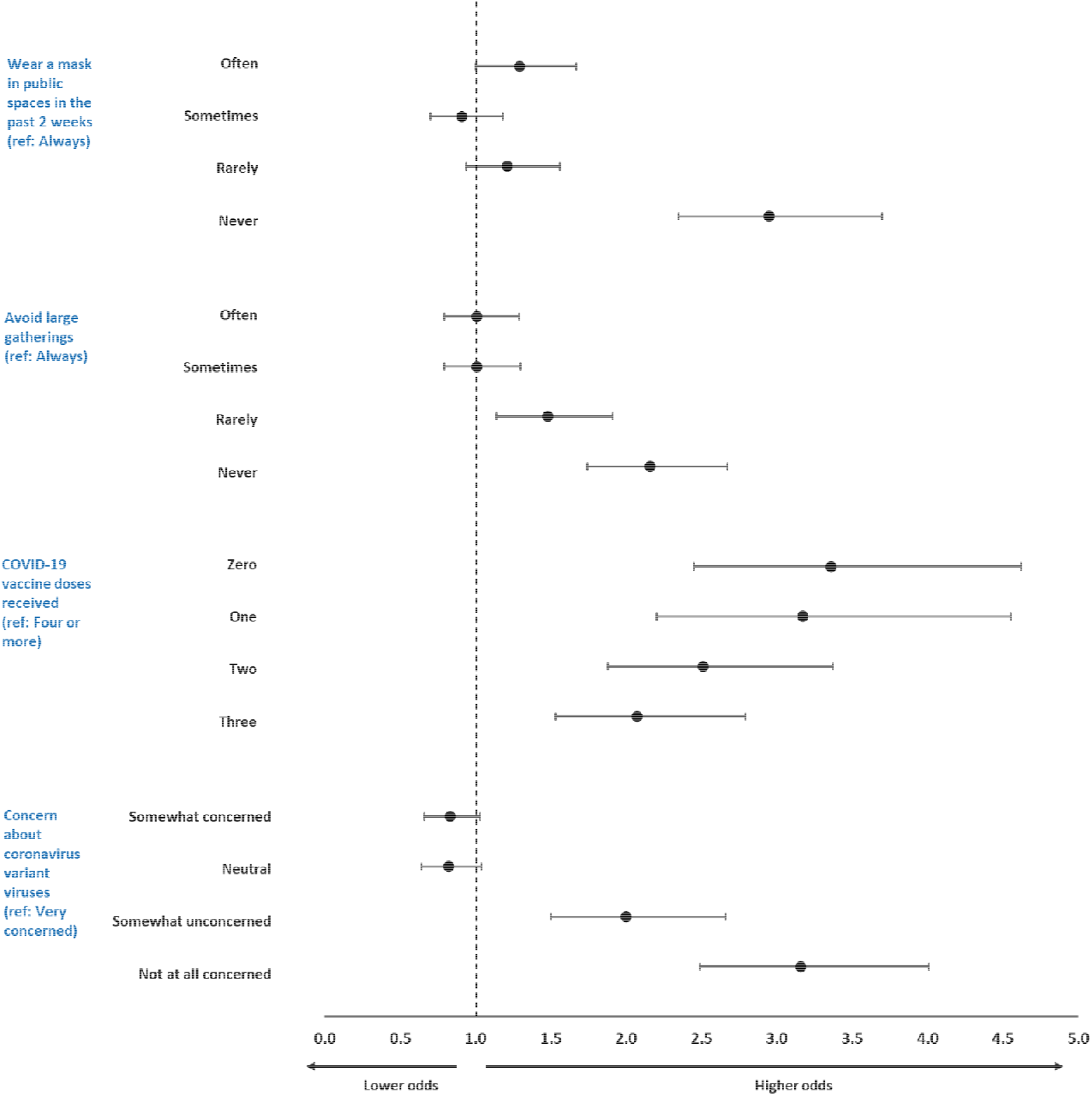
Adjusted Odds Ratios of Having Agreed that the Pandemic was Over by COVID-19-Related Characteristics, September to October 2022.

## Discussion

Only 1 in 3 US adults in a large scale, demographically representative survey sample agreed “the [COVID-19] pandemic is over” in September to October 2022. In contrast, 43% of adults disagreed that the pandemic was over, while the remaining 24% were neutral about the statement. Agreement with the pandemic-over statement was most strongly associated with having received fewer COVID-19 vaccines, lesser concern about SARS-CoV-2 variant viruses, and less frequent engagement in COVID-19 preventive behaviors such as mask usage in public spaces, as well as increasingly conservative political ideology, roles as unpaid caregivers of both children and adults, younger age, male sex, and significant disabilities. These findings demonstrate how divided public sentiments are about the state of the COVID-19 pandemic, and reveal associated COVID-19-related attitudes and behaviors that could compromise mitigation efforts, as well as associated demographic characteristics that could inform tailored messaging strategies.

Inconsistent messaging about the COVID-19 outbreak has been a persistent challenge over the course of the pandemic. For example, through February and March 2020, many US public health leaders and the WHO advised the public against use of face masks to reduce SARS-CoV-2 transmission, even stating that they could increase risk of infection, and reported that asymptomatic transmission of SARS-CoV-2 was rare.^**22–24**^ In April 2020, the CDC recommended public face mask use for the first time, a stance strengthened by convincing evidence of the effectiveness of masks in reducing SARS-CoV-2 infections and COVID-19 hospitalizations and deaths.^**25–29**^ However, revision of the guidance on masking plagued widespread adherence with this affordable, noninvasive, effective behavior to reduce SARS-CoV-2 transmission due in part to the complete reversal of prior guidance and continued belief that masks were ineffective by a portion of the public.^**30,31**^ Similarly, in May 2021 the CDC lifted mask and social distancing recommendations for fully vaccinated individuals, citing extremely low risks of infection.^**32**^ The guidance suggested low breakthrough SARS-CoV-2 infections among vaccinated persons, even indicating that fully vaccinated persons need not test following exposures, though breakthrough infections associated with severe COVID-19 illness have occurred at non-negligible rates.^**33**^ Mixed messages have continued to present a challenge for public health leaders for guidance regarding the return to in-person school and work, aerosolized transmission of coronaviruses, and SARS-CoV-2 variant viruses.^**34**^

Most recently, there has been controversy regarding revised CDC guidelines for minimizing the impact of COVID-19, published in August 2022.^**35**^ The guidelines, which do not mention the word pandemic, have been interpreted as transferring responsibility to individuals to monitor the pandemic and take actions to protect themselves from severe COVID-19 outcomes^**36**^ at a time when COVID-19 media coverage has declined.^**37**^ Moreover, the guidelines mention but do not discuss Long COVID, which can affect people across the spectrum of acute COVID-19 illness, including initially asymptomatic infections,^**38**^ leading to considerable healthcare costs.^**39,40**^ Despite evidence that risk of Long COVID increases with reinfections regardless of vaccination status,^**41**^ pandemic-related public health messaging has not yet focused on minimizing risk of Long COVID, instead focusing on strategies to mitigate severe outcomes in the acute stage of illness.

The findings of the current study are also consistent with those of a study about perceived local COVID-19 transmission levels,^**20**^ as both studies found that individual behaviors aligned with peoples’ perceptions of important pandemic-related information. People who agreed the pandemic was over and those who did not accurately perceive local COVID-19 transmission levels as high had received fewer COVID-19 vaccines, conveyed less concern about SARS-CoV-2 variant viruses, and reported less frequent engagement in COVID-19 preventive behaviors compared with other adults. The extent to which individuals are no longer seeking COVID-19-related information or the information is not readily available is unclear. Given the association between these beliefs and non-engagement in COVID-19-related prevention behaviors, public health communication efforts to fully inform these groups are needed.

The findings of demographic characteristics associated with agreement with the pandemic-over statement have important implications for tailored public health messaging about the pandemic. Some associations with agreement, such as more conservative political ideology, were anticipated given political polarization during the pandemic, as more conservative ideology has consistently been associated with lower levels of concern about and attention to the pandemic and less frequent engagement in COVID-19 prevention efforts.^**42**^

However, the findings that agreement with the pandemic-over statement was associated with significant disabilities and roles as unpaid caregivers of both children and adults were surprising. People with disabilities are at increased risk of severe COVID-19 outcomes, as highlighted in the CDC’s guidance published in August 2022.^**35**^ Indeed, COVID-19-associated hospitalization rates among disability-eligible Medicare beneficiaries are approximately 50% higher than rates among age-eligible beneficiaries.^**43**^ For other major risk factors, such as age and health status, responses to the pandemic-over statement reflected risk of severe COVID-19 outcomes, as older adults and people with fair or poor vs excellent health more commonly disagreed that the pandemic was over. Perhaps not surprisingly, caregivers of children and adults, who’s caregiving often puts them at higher risk of SARS-CoV-2 exposures, more commonly agreed that the pandemic was over. For caregivers of children specifically, perceptions could also be influenced by seasonal outbreaks of influenza and respiratory syncytial virus detracting attention away from COVID-19. Further investigation into the factors underlying perceptions about the pandemic within these groups is warranted.

Strengths of this analysis include use of a large, demographically representative sample of US adults and comprehensive characterization of respondents. There are limitations, including English-language-only, internet-based surveys that were administered using non-probabilistic sampling methods, limiting the generalizability of the findings; however, demographic quota sampling and survey weighting were applied to improve demographic representativeness. Additionally, associative findings from multivariable models do not reflect directionality or causality.

In conclusion, based on a large scale, demographically representative sample, we found that as of late September to mid-October 2022, only 1 in 3 US adults agreed with the statement “the pandemic is over” and that 43% of adults disagreed with that statement. Adults who agreed with the pandemic-over statement had higher odds of being unvaccinated or under-vaccinated against COVID-19, were less concerned about SARS-CoV-2 variant viruses and were less engaged in behaviors to reduce COVID-19 transmission. We also identified demographic groups to prioritize for tailored public health messaging about the pandemic status, including people with more conservative political ideology, with significant disabilities, in roles as unpaid caregivers of both children and adults, and of younger age or male sex. Continued assessment of public perceptions about the state of the pandemic is warranted as we enter Year 4 of the COVID-19 pandemic.^**44**^

## Data Availability

All data produced in the present study are available upon reasonable request to the authors and institutional agreement.

## Additional Information

### Author Contributions

M. É. Czeisler had full access to all of the data in the study and takes responsibility for the integrity of the data and the accuracy of the data analysis.

### Concept and design

M. É. Czeisler, Weaver, C. A. Czeisler.

### Acquisition, analysis, or interpretation of data

M. É. Czeisler, Weaver.

### Drafting of the manuscript

M. É. Czeisler.

### Critical revision of the manuscript for important intellectual content

All authors.

### Statistical analysis

M. É. Czeisler.

### Obtained funding

M. É. Czeisler, Weaver, Lane, Howard, Rajaratnam, C. A. Czeisler.

### Administrative, technical, or material support

Weaver, Lane, C. A. Czeisler.

### Disclaimer

The findings and conclusions in this report are those of the authors and do not necessarily represent the official position of the Centers for Disease Control and Prevention.

