## Supplement for "US Public Opinion as to Whether “The Pandemic is Over,” September to October 2022"

Title:

Study Approval and Reporting

This activity was reviewed by the US Centers for Disease Control and Prevention (CDC) and was conducted consistent with applicable federal law and CDC policy (i.e., 45 C.F.R. part 46, 21 C.F.R. part 56; 42 U.S.C. Sect. 241(d); 5 U.S.C. Sect. 552a; 44 U.S.C. Sect. 3501 et seq.). This study followed the American Association for Public Opinion Research (AAPOR) reporting guideline.

Demographic Characteristics

Sex assigned at birth was assessed with response options of Male, Female, Intersex, Refused or prefer not to say, and I don’t know. Participants who reported male or female sex were included to align with US Census response options for survey weighting. Age was assessed with continuous response options and defined as a categorical variable with 4 groups: 18 to 24 years, 25 to 44 years, 45 to 64 years, or 65 years and over. Combined race and ethnicity were assessed among survey respondents with separate questions and options defined by the investigators based on US Census classifications. The race and ethnicity questions follow.

1. What is your race? (Select all that apply)
   a. American Indian or Alaskan Native
   b. Asian
   c. Black or African American
   d. Native Hawaiian or other Pacific Islander
   e. White
   f. Other

Please use the categories that most reflect your recognition in the community for purposes of reporting mixed racial and/or ethnic origins.

American Indian or Alaskan Native: a person having origins in any of the original peoples of North, Central, or South America, and maintains tribal affiliations or community attachment.

Asian: A person having origins in any of the original peoples of the Far East, Southeast Asia, or the Indian subcontinent including, for example, Cambodia, China, India, Japan, Korea, Malaysia, Pakistan, the Philippine Islands, Thailand, and Vietnam.

Black or African American: A person having origins in any of the black racial groups of Africa.

Native Hawaiian or Pacific Islander: A person having origins in any of the original peoples of Hawaii, Guam, Samoa, or other Pacific Islands.

White: A person having origins in any of the original peoples of Europe, North Africa, or the Middle East.

2. What is your ethnicity? (Select one)
   a. Hispanic or Latino
   b. Not Hispanic or Latino

Hispanic or Latino: A person of Cuban, Mexican, Puerto Rican, South or Central American, or other Spanish culture or origin regardless of race.

For this analysis, race and ethnicity were combined into the following categories: White, non-Hispanic; Black, non-Hispanic; Asian, non-Hispanic; Multiple races or Other race, non-Hispanic; or Hispanic, any race or races. Highest education attainment was defined as a categorical variable with 3 groups: High school degree or less, College or some college, or More than bachelor’s degree. Experience with disability was defined based on responses to the 6-item Washington Group Short Set of Disability Questions, used to assess whether people have difficulty performing basic universal activities such as walking, seeing, hearing, cognition, self-care and communication.^1^ Participants were assigned groups according to their most significant limitation (ie, if they could not do one or more tasks at all, they were assigned to that group; if they had no difficulty with any tasks, they were assigned to that group), such that experience with disability was a categorical variable with 4 groups: No difficulty with any tasks, Some difficulty with tasks, A lot of difficulty with tasks, or Cannot do some tasks at all. Self-rated health status was defined based on responses to the first question in the 36-Item Short Form Health Survey,^2,3^ with response options of Excellent, Very good, Good, Fair, or Poor. Unpaid caregiver roles were defined as a categorical variable with 4 groups: Unpaid caregiver of adults aged ≥18 years only, Unpaid caregiver of children aged <18 years only, Unpaid caregiver of persons in both age groups, or None (ie, not an unpaid caregiver). US Census region was categorized based on the state of residence reported by participants, defined as a categorical variable with 4 groups: Northeast, Midwest, South, or West. Political ideology was defined as a categorical variable with 5 groups: Very liberal, Slightly liberal, Neither liberal nor conservative, Slightly conservative, or Very conservative.

COVID-19-related Characteristics

Frequency of mask usage in public spaces in the 2 weeks preceding the survey was defined as a categorical variable with 6 groups: Always, Often, Sometimes, Rarely, Never, or I did not go to a public place. Frequency of avoidance of large gatherings in the 2 weeks preceding the survey was defined as a categorical variable with 6 groups: Always, Often, Sometimes, Rarely, or Never. COVID-19 vaccine doses received were defined as a categorical variable with 5 groups: Zero, One, Two, Three, or Four or more. Concern about coronavirus variant viruses was defined as a categorical variable with 5 groups: Very concerned, Somewhat concerned, Neutral, Somewhat unconcerned, and Not at all concerned.
